# Effectiveness and mechanisms of a multimodal treatment for low back pain: a pragmatic matched cohort study

**DOI:** 10.1101/2022.09.28.22280380

**Authors:** Robin Schäfer, Daniel Niederer, Claudia Levenig, Monika Hasenbring, Thomas Tas, Daniela Fett, Katharina Trompeter, Thore Haag, Christian Schneider, Philipp Floessel, Heidrun Beck, Marcus Schiltenwolf, Pia-Maria Wippert, Tilman Engel, Frank Mayer, Petra Platen

## Abstract

**Objective:** To investigate the effectiveness and mechanisms of a multimodal treatment including perturbation exercise.

**Methods:** A matched cohort study was conducted. The intervention consists of a 12-week back pain prevention course with perturbation exercise and education embedded in primary health care according to German social law. Participants from the intervention group had chronic or recurrent low back pain with heterogenous but on average rather low pain and chronification. Control groups (usual care) were matched from a multi-center RCT. Outcomes were pain, disability, isokinetic trunk strength and balance. Bayesian regression models were used to estimate the Average Treatment effect on the Treated (ATT). Further, sub-group and mediation analyses within the intervention group using the biopsychological avoidance-endurance model were conducted. Median values with highest posterior density intervals (HPDI) from baseline-adjusted analyses are presented.

**Results:** Over 12 weeks, intervention and control (n = 128 each) experienced a similar decrease in pain and disability, which led to negligible ATTs for pain (−0.3 (HPDI95% [-4.3, 3.4]) and disability (−0.2 (HPDI95% [-4, 3.7])). Changes in functional parameters (n=18) showed small effects in favor of the intervention group, in particular for monopedal stances (standardized mean difference: -0.5 HPDI95% [-0.79, -0.21]). Depression was higher in drop-outs and decreases in pain/disability were associated with decreases in depression. Distress-endurance subgroups experienced higher baseline pain and disability and showed the highest reductions in both parameters upon completion of the intervention.

**Conclusion:** Perturbation exercise with education yielded only small treatment effects in a heterogenous population with rather mild symptoms. Targeting distress-endurance subgroups with a multimodal treatment approach is probably an effective strategy in treatment tailoring.

## Introduction

The burden of low back pain (LBP) is high [9] and treatment effect sizes are usually only small to medium [21]. One reason for that low effect is that primary pathologies and optimal treatments for relevant subgroups in the LBP domain can barely be found [21, 40]. Therefore, the use of a biopsychosocial model for explaining LBP (especially at the chronic stage or chronification) and guiding treatment (e.g. multidisciplinary/-modal) is emphasized [4, 22]. While education and exercise are almost consistently recommended as first-line treatment [13], the use of a costly and time-intensive multidisciplinary approach should be tailored (e.g. for chronic LBP with significant psychosocial contribution) [8, 22].

The biopsychosocial avoidance-endurance model from Hasenbring et al. [15] aims to describe the chronification of pain by two maladaptive patterns: *avoidance* and *endurance* (*overdoing)* [6]. Endurance-related responses can be further distinguished to distress-endurance (pain-related thought suppression, depressive mood) and eustress-endurance (focused distraction, positive mood) which both result in task-persistence and physical overuse [18]. The fear-avoidance response includes pain-related avoidance of activity and catastrophizing. The fear-avoidance and endurance response result in physical dis-/overuse [18] and therefore negatively affect pain intensity, disability, pain severity and quality of life [28]. Consequently, *activity pacing* (adaptive response) is proposed as an optimal reaction to the onset of pain. High levels of activity pacing and low levels of avoidance were associated with better outcomes for physical functioning [6]. The distress-endurance might be the most critical subgroup regarding experienced pain and disability [12, 16, 43]. However, we do not know yet if those subgroups react differently to a multimodal treatment including exercise. Further, it is unknown whether changes in underlying subscales according to the avoidance-endurance model mediate changes in pain and disability outcomes.

In this study, we aim to examine the effectiveness of a multimodal treatment (sensorimotor exercise with perturbation + education) in primary, preventive care. setting [7, 11, 37]. The effects are compared to a matched control group from a multi-center RCT, which evaluated the exercise component’s *efficacy* [31, 32]. Further, we want to examine moderating (i.e., for whom is the treatment effective?) and mediating (i.e, how does the treatment work?) factors by integrating the avoidance-endurance model. Specifically, we hypothesize that changes towards adaptive behavior lead to beneficial changes in pain and disability.

## Methods

### Study Design and Ethics

We carried out a pragmatic, prospective cohort study with matched pairs. The study was conducted in agreement with the declaration of Helsinki, approved by the local ethics committee of the Faculty of Sport Science of Ruhr-University Bochum (EKS V 10/2017). All participants gave their written informed consent after receiving written and oral information about the study. The trial was retrospectively registered in the German Clinical Trial Register (DRKS00030389). The study report follows the guidelines CERT [38] and the CONSORT extension for pragmatic trials [49].

### Participants

#### Intervention group

Participants were recruited from a specific back pain prevention program. The program was offered by their health insurance company (AOK Nordwest) free of charge. Such prevention programs are part of primary health care as they are anchored in the German social law (SGB V $20a). The participants either self-registered for the program or were offered this program in a personal health consultation by employees of the health insurance company or the executing coaches.

#### Control group

The control group was matched from a cohort of a multi-center study conducted within our study network [32]. The control group received no intervention. However, they received a more frequent and extensive diagnostic assessment than the intervention group within the given timeframe of 12 weeks. The intervention and control group from this multi-center study were not prohibited from proceeding with their usual care or receiving additional treatments (e.g. physiotherapy) [32].

#### Inclusion and exclusion criteria

Inclusion criteria for both groups were: age 18-65, at least one episode (≥ 4 days) of nonspecific back pain in the last 12 months, ability to understand the extent and meaning of the study and to answer a questionnaire without help [32]. The exclusion criteria were pregnancy, acute pain during the past seven days, being unable to stand upright, not able to give outcome sick leave information, or showing signs of acute risk factors referred to as “red flags” (inflammatory, traumatic or systematic processes).

### Intervention

The intervention consisted of a 12-week multimodal group intervention program including sensorimotor exercises with perturbations, and theoretical education. The exercise component from a multi-center study [32] was modified to be delivered in a group instead of personal coaching and an education component was added. The intervention took place at multiple locations and was, thus, given by different coaches. Prior to the study, all coaches received a full-day workshop and were educated in theory and practice about the content of the intervention program by a multidisciplinary team of our research network.

The exercise components comprised four core exercises (plank, side-plank, single-leg stance, rowing) which were varied in stability and strength demands [32, 36]. The participants of the course were supposed to exercise one session per week á 60 minutes in the group supervised by the coaches (center-based) and were instructed to do two additional weekly sessions á 30 minutes at home (home-based). The home-based training was not documented by the participants or the coaches.

The education components as part of the center-based sessions included pain neurophysiology, pain coping strategies according to the avoidance-endurance model [16, 18] and other relevant topics in the context of back pain (e.g. physical activity, activity pacing, stress & relaxation, risk factors, sensorimotor exercise).

According to the CERT guideline, further details are presented in the supplement (https://osf.io/c84rt).

### Outcomes

The main outcomes pain intensity and pain disability were measured with the Chronic Pain Grade (CPG) questionnaire in the intervention and control group. Within a subset of the intervention and the control group isokinetic trunk strength (extension/flexion) and balance (postural sway) were assessed. Finally, and only within the intervention group, avoidance-endurance related parameters (e.g., depression, pain persistence) and subgroup classifications based on the avoidance-endurance model [17] were evaluated.

#### Pain & Disability

The 7-item Chronic Pain Grade (CPG) questionnaire by von Korff et al [25] was used to assess pain intensity and pain disability on a 10-point Likert Sale, which can be scored within a characteristic pain intensity and subjective pain disability scale. Furthermore, different grades of chronification can be calculated as described in the original literature [25].

#### Strength & Balance

The maximum isokinetic trunk strength (extension and flexion) was tested with the IsoMed 2000 Back Module (D&R GmbH, Germany). After a warm-up phase of 30 submaximal repetitions, the test consisted of five maximum repetitions of 20° extension and 30° flexion in a sitting position. The movement speed was constant at 60°/s. The average value of the three highest repetitions was used as outcome [Nm] for flexion and extension.

Balance was tested in bipedal and monopedal stances (left and right) over 30 s. The trace of the center of pressure [mm] was calculated as an outcome parameter, where the sum of both monopedal stances was used as outcome. Stances were standardized (hands gripping the hip, barefoot, visual fixation in 2-meter distance). The tests were performed on different force platforms for the intervention (Balance Trainer BTG4, Hur Labs, Finnland) and the control group (CSMi Computer Sports Medicine Inc., Stoughton, MA, USA). We performed a direct comparison of these devices and both instruments yielded valid results for the postural sway [3]. However, to account for heterogeneity, pre and post scores were standardized by the mean and standard deviation of the pre-test within each group (intervention/control).

#### Depression

Depression was measured by the German version of the Beck Depression Inventory for Primary Care (BDI-PC) [33] and two extra items (“loss of pleasure” and “incapacity to decide”) from the long BDI [2, 26]. The BDI-PC consists of 7 items, where the sum of those builds the subscale for depression ranging from 0 (no depression) to 21 (highest possible depression).

#### Avoidance-Endurance

Psychosocial responses were measured by the Avoidance-Endurance Questionnaire (AEQ) [17]. The questionnaire consists of 28 items. The pain persistence and humor/distraction subscales are part of the behavioral endurance scale of the AEQ. Further scales used in this study were thought suppression and hopelessness.

#### Subgroup classification

The classification of the avoidance-endurance subgroups was based on 1) the pain persistence scale (7 items) from the AEQ and the BDI-PC (7 items) and 2) the pain persistence scale and two items from the extended BDI version [43, 48]. In both procedures, pain persistence values equal or higher than 3 led to an endurance classification (either distress- or eustress-endurance) and otherwise to adaptive/avoidance response. For 1), depression values equal to or higher than 2 led to a distress-endurance or fear-avoidance classification and otherwise to an eustress-endurance or adaptive classification. For 2), the same classification was applied if both depression items were rated 1 or higher [41].

### Statistical Analyses

The results should be considered rather explorative and hypothesis-generating than confirmative. Therefore, no significance testing was performed. The effects were described using *confidence* or *highest posterior density intervals*, which we aim to interpret as compatibility intervals [1]. All analyses were conducted in R [34].

#### Matching & Treatment Effects

Two control groups were matched from a multi-center RCT to calculate treatment effects for pain and disability (Cohort 1) and functional outcomes (Cohort 2). The matching was done by calculating the (multivariate) Mahalanobis Distance between each observation [24]. A genetic matching algorithm was used, which aims to pick 1:1 matches by minimizing an imbalance function [10]. Baseline values from the respective outcomes, sex and age were finally used as covariates for matching. Matching was performed in R using the MatchIt package [20].

Bayesian regression models were used to estimate the ATT (Average Treatment Effect on Treated) for each outcome (pain, disability, strength, balance) [14]. Three outcome models (a baseline-adjusted, a change score and a full covariate-adjusted model) incorporating weights from matching were fitted. The models were fitted with the brms package [5] using a weakly informative prior for pain/disability (normally distributed change score with mean = 0 and standard deviation = 10), strength (mean = 0, standard deviation = 10, unit: Nm) and balance (mean = 0, standard deviation = 1, unit: SMD). The posterior distribution of the ATT was then obtained for each model using g-computation [39]. Further, the model-averaged posterior for all three models was calculated. The *highest posterior density intervals* (e. g. 95%) from the model-averaged posterior distributions were visualized and reported. Further, we analyzed discrete changes in *avoidance-endurance* classifications and changes in the underlying subscales mostly on a descriptive basis.

#### Sub-group analyses within the intervention group

Sub-group analyses for the outcomes characteristic pain intensity and subjective pain disability were performed for avoidance-endurance subgroups and chronification grades. Simple linear regression models with the change of pain intensity and pain disability as outcome and subgroup as a predictor were calculated. Baseline-adjusted models were used for sensitivity analysis.

#### Mediation within the intervention group

We further evaluated if the treatment works via changes in subscales of the avoidance-endurance model affecting characteristic pain intensity and subjective pain disability as outcomes using the within-participant mediation model from Montoya & Hayes [29]. The model was adopted and enhanced to multiple mediators (depression, hopelessness, thought suppression, pain persistence, distraction) and multiple outcomes (pain intensity, pain disability) (Figure 1). Direct and indirect (mediated) treatment effects were calculated for each subscale and each outcome. The grand mean centered parametrization from Montoya and Hayes [29] was used to meaningfully interpret the results. The models were fitted via the Bayesian regression package brms [5]. The full posterior distribution is presented for direct, indirect and total effects.

**Figure 1.**
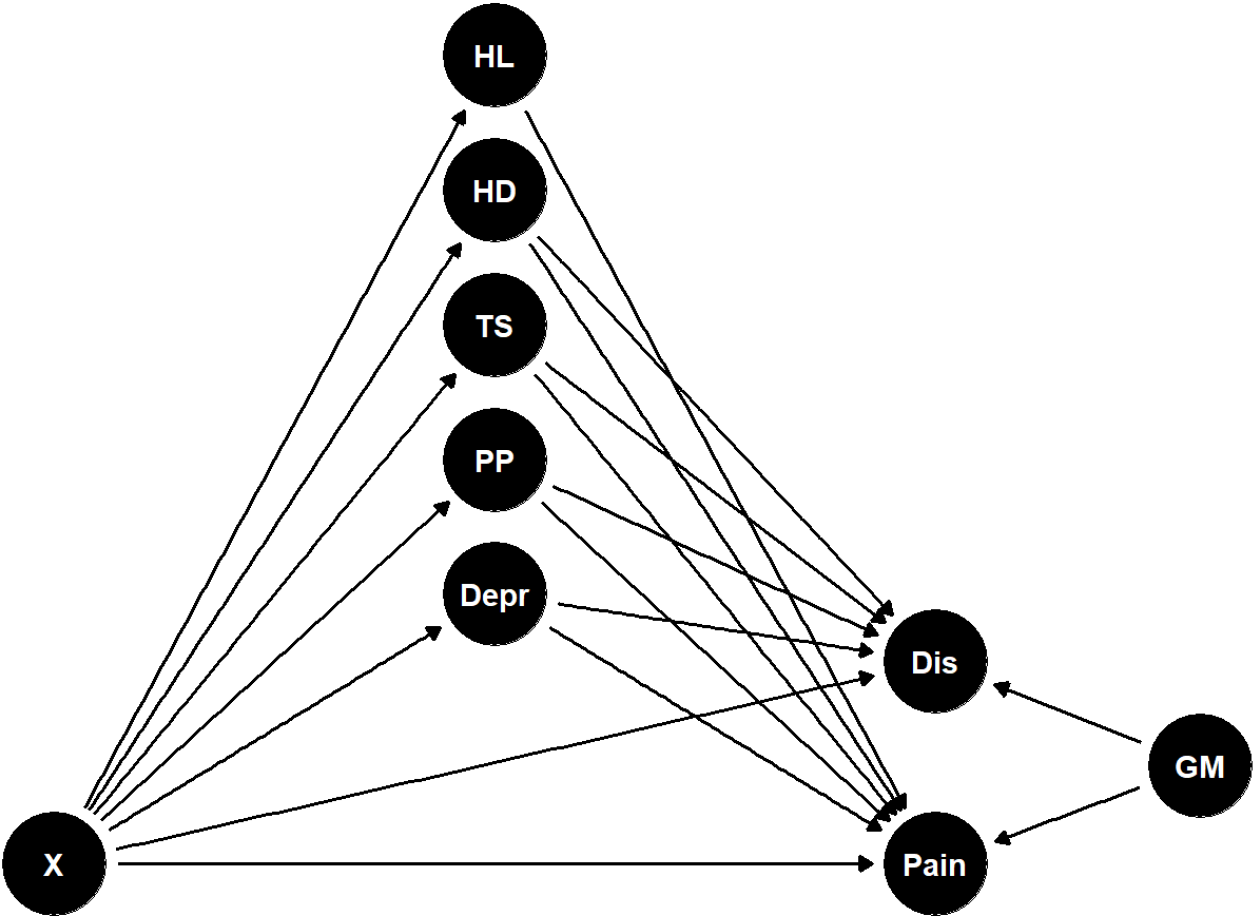
Causal Graph of the one-condition within-participant mediation analysis with multiple serial mediators and 2 outcomes adapted from Montoya & Hayes [33]. The nodes for mediators and outcomes represent pre-post differences. X = treatment (intervention only), Mediators: Depr = depression, PP = pain persistence, TS = thought suppression, HD = humor/distraction, HL = hopelessness. Outcomes: Dis = disability, Pain = pain intensity, Parametrization: GM = grand mean

## Results

Figure 2 shows the participant flow for both groups (intervention and control) and both matching procedures (pain and functional outcomes). Dropout during the intervention period was high (60.1%, n = 190 who did not show up at the last course session or did not answer the questionnaire). The trainers reported no adverse events. The baseline values of participants who dropped out tended to be higher for psychological subscales (e.g., hopelessness, catastrophizing, thought suppression) and pain/disability.

**Figure 2:**
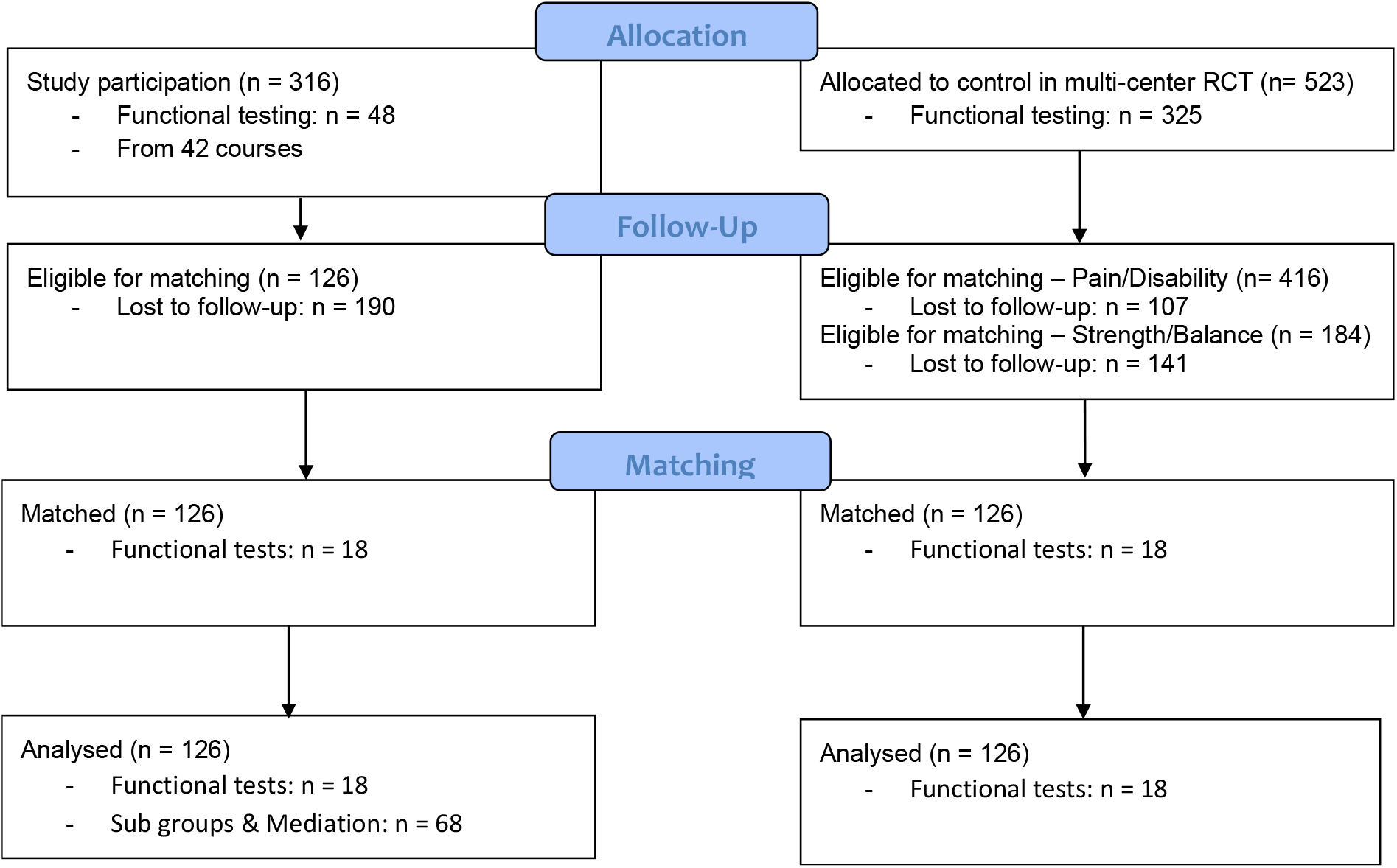
CONSORT flow diagram.

### Balance assessment

Covariate balance for demographics and baseline outcomes in both matching procedures was reasonable. Small to moderate baseline imbalance was observed for age (Standardized Mean Difference (SMD) = 0.11 and pain (SD = 0.08) when matching for pain/disability. When matching for functional outcomes, baseline imbalance was SMD = -0.12 for flexion, SMD = 0.12 for extension and SMD = 0.13 for age. Mean values for all covariates and matching procedures are presented in Table 1.

**Table 1:**
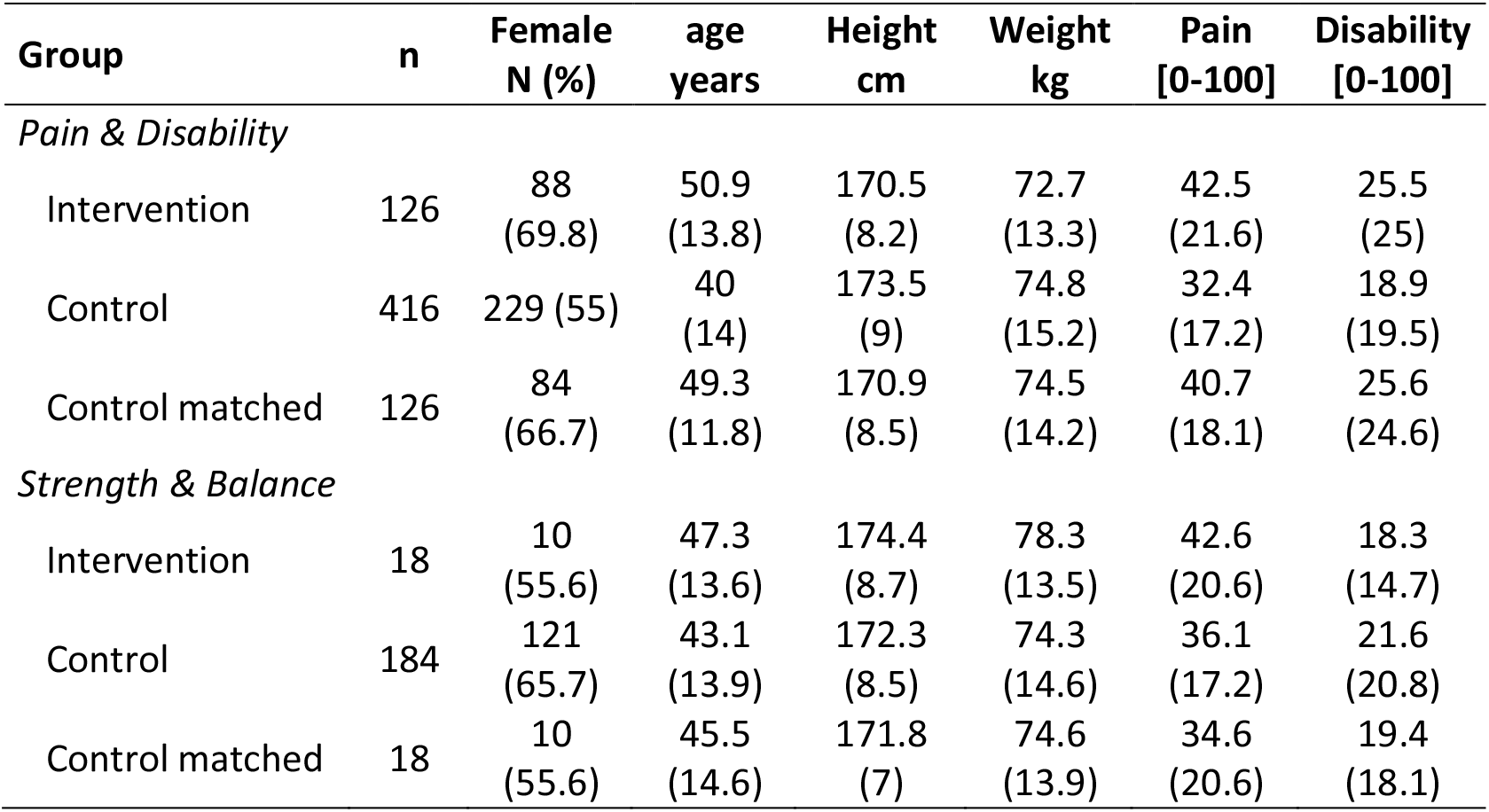
Covariates of the intervention and control group before and after matching

| Group | n | Female<br>N (%) | age<br>years | Height<br>cm | Weight<br>kg | Pain<br>[0-100] | Disability<br>[0-100] |
| --- | --- | --- | --- | --- | --- | --- | --- |
| *Pain & Disability* |  |  |  |  |  |  |  |
| Intervention | 126 | 88<br>(69.8) | 50.9<br>(13.8) | 170.5<br>(8.2) | 72.7<br>(13.3) | 42.5<br>(21.6) | 25.5<br>(25) |
| Control | 416 | 229 (55) | 40<br>(14) | 173.5<br>(9) | 74.8<br>(15.2) | 32.4<br>(17.2) | 18.9<br>(19.5) |
| Control matched | 126 | 84<br>(66.7) | 49.3<br>(11.8) | 170.9<br>(8.5) | 74.5<br>(14.2) | 40.7<br>(18.1) | 25.6<br>(24.6) |
| *Strength & Balance* |  |  |  |  |  |  |  |
| Intervention | 18 | 10<br>(55.6) | 47.3<br>(13.6) | 174.4<br>(8.7) | 78.3<br>(13.5) | 42.6<br>(20.6) | 18.3<br>(14.7) |
| Control | 184 | 121<br>(65.7) | 43.1<br>(13.9) | 172.3<br>(8.5) | 74.3<br>(14.6) | 36.1<br>(17.2) | 21.6<br>(20.8) |
| Control matched | 18 | 10<br>(55.6) | 45.5<br>(14.6) | 171.8<br>(7) | 74.6<br>(13.9) | 34.6<br>(20.6) | 19.4<br>(18.1) |

### Characteristic Pain Intensity & Pain Disability

The model-averaged ATT (average treatment effect of the treated) was -0.3 (HPDI95% [-4.3, 3.4]) for pain intensity and -0.2 (HPDI95% [-4, 3.7]) for pain disability, where negative values favor the intervention. Considering the range of the scales (0 to 100) and compatibility intervals smaller than 5, the ATT would indicate no substantial treatment effect (see Figure 3). However, there was an average decrease in both groups for pain intensity (intervention: 8.0 SD: 18.7, control: 7.0 SD: 18.5) and pain disability (intervention: 6.5 SD: 21.5, control: 6.7 SD: 20.7).

**Figure 3.**
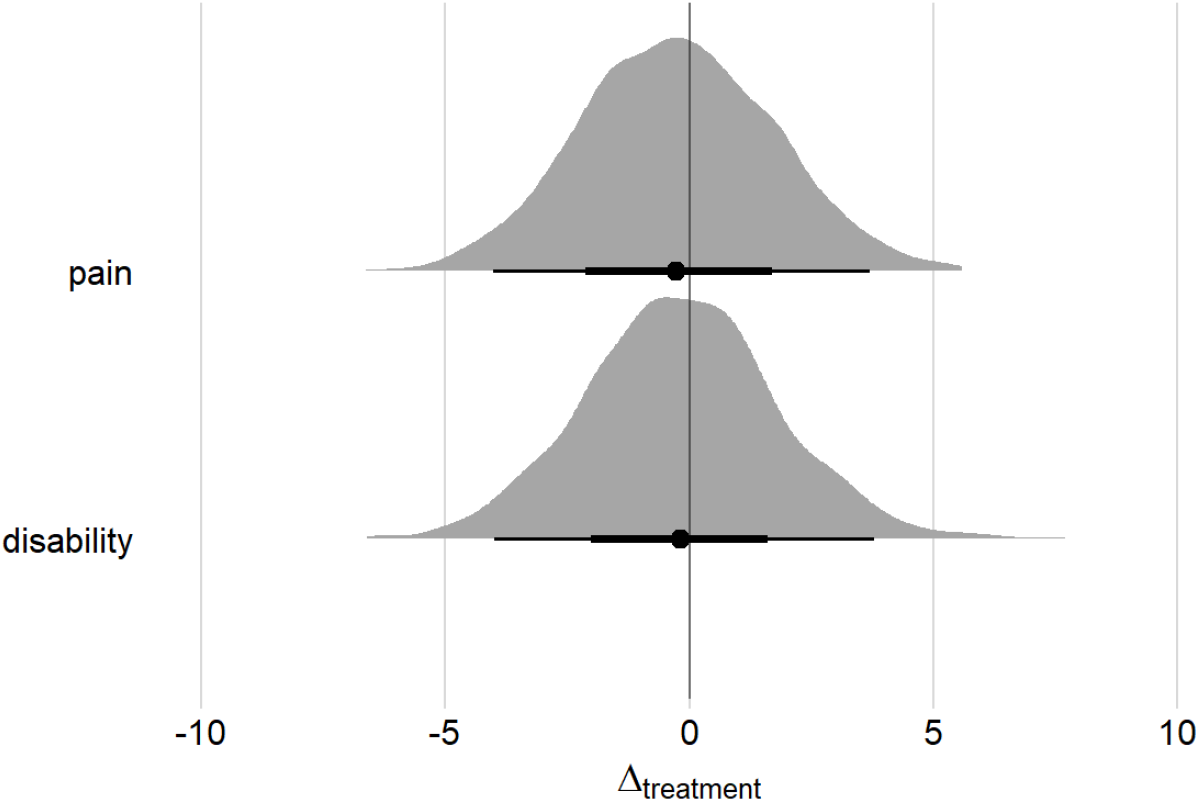
Posterior distributions for the model-averaged ATT (Average Treatment effect on Treated) for characteristic pain and pain disability– n = 126. The distributions were stacked from 3 outcome models (ANCOVA, change score, covariate-adjusted). Dots show the median of the posterior distribution and bold/thin whiskers represent 95/50% HPDI (highest posterior density interval)

**Figure 4.**
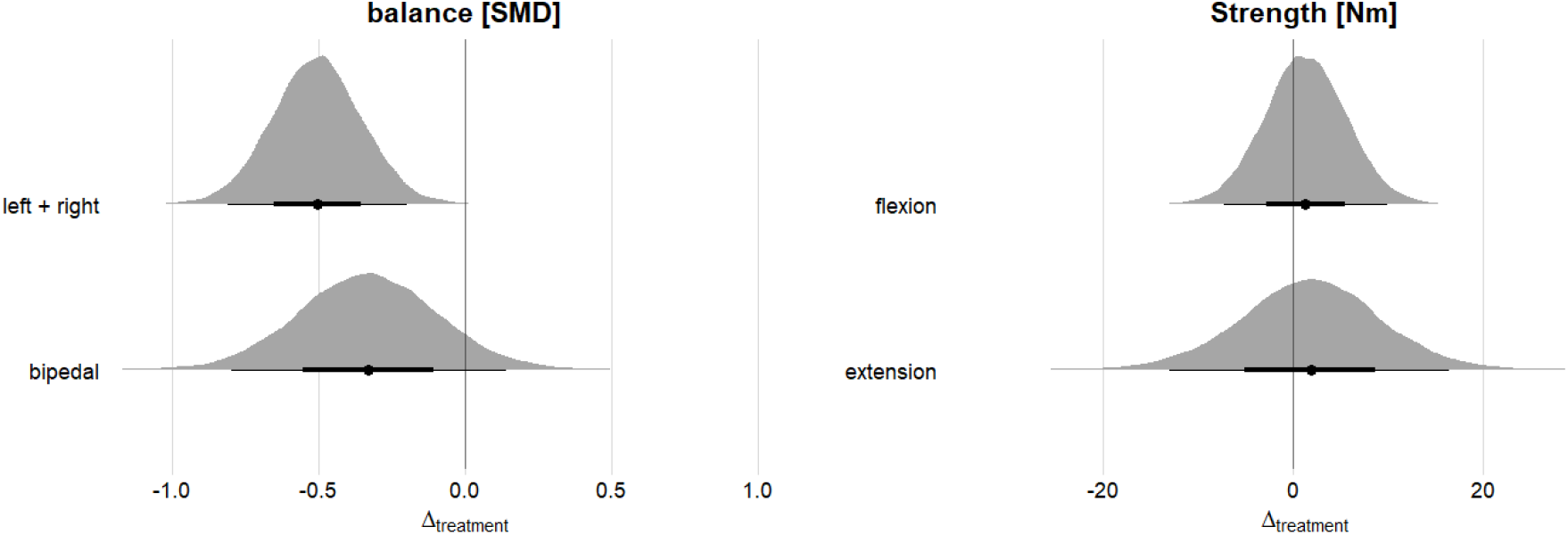
Posterior distributions for the model-averaged ATT (Average Treatment effect on Treated) for the functional outcomes balance and strength – n = 18. The distributions were stacked from 3 outcome models (ANCOVA, change score, covariate-adjusted). Dots show the median of the posterior distribution and bold/thin whiskers represent 95/50% HPDI (highest posterior density interval)

### Strength & Balance

The model-averaged ATT indicates mostly favorable adaptations in strength and balance for the treatment group, where negative values for balance and positive values for strength favor the intervention. Postural sway reductions compared to control were larger in monopedal stance (sum of left and right; SMD = -0.5 HPDI95% [-0.79, -0.21]) than those in bipedal stances (−0.32 HPDI95% [-0.79, 0.11]). The ATT was similar for trunk extension (2.1 Nm HPDI95% [-13.2, 16.4]) and flexion (1.1 Nm HPDI95% [-7.3, 10.0]). However, the treatment effects for strength compared to balance were more uncertain (lower and wider curves) and smaller on average.

### Avoidance-Endurance Parameters

In the intervention group (n=68), the mean reductions for pain persistence (scale: 0 to 6) and depression (scale: 0 to 9) were 0.1 (SD: 0.9) and 0.9 (SD: 2.4), respectively. These changes were driven by the participants who had high baseline values. Given that the subgroups were determined by depression and pain persistence, this led to substantial positive changes in subgroup classifications after the treatment (see Table 2). The favorable adaptive classification increased from 16 to 30 and the maladaptive distress-endurance type was reduced from 22 to 12.

**Table 2.** Avoidance-Endurance classifications before and after the treatment in the intervention group (n=68). The classification is based on *pain persistence* and *depression* subscales.

|  | Adaptive | Distress-Endurance | Eustress-Endurance | Fear-Avoidance |
| --- | --- | --- | --- | --- |
| Pre | 16 (23.5%) | 22 (32.4%) | 20 (29.4%) | 10 (14.7%) |
| Post | 30 (44.1%) | 12 (17.6%) | 19 (27.9%) | 7 (10.3%) |

### Sub-group analyses – for whom is the treatment effective?

Evaluating the treatment group only (n=68), characteristic pain intensity and subjective pain disability reductions were highest for the distress endurance subgroup (see Table 3). However, adjusting the analysis for baseline values diminished the outcome for distress-endurance. In this subsample, reductions were slightly higher for pain intensity (−9.9 SD: 17.5) and pain disability (−9 SD: 23.4) compared to the complete matched cohort (n=126).

**Table 3.**
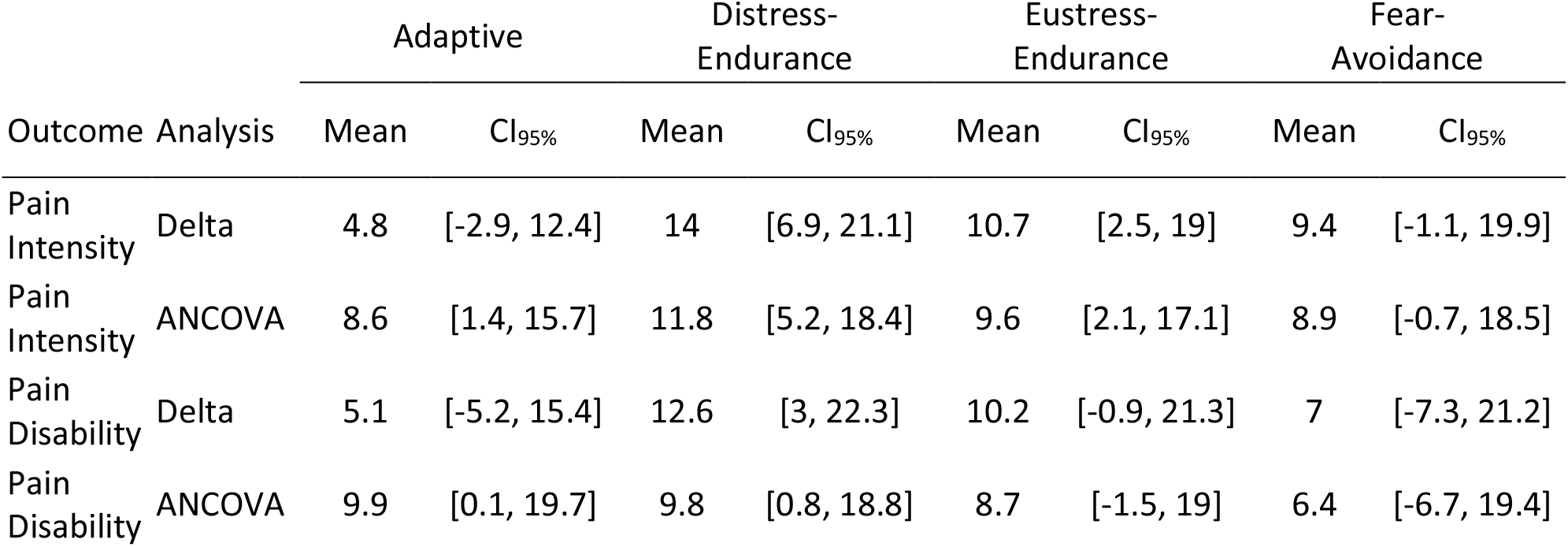
Moderation analysis for characteristic pain intensity and subjective pain disability reductions stratified by avoidance-endurance subgroups at baseline in the intervention group (n=68)

|  |  | Adaptive |  | Distress-Endurance |  | Eustress-Endurance |  | Fear-Avoidance |  |
| --- | --- | --- | --- | --- | --- | --- | --- | --- | --- |
| Outcome | Analysis | Mean | CI<sub>95%</sub> | Mean | CI<sub>95%</sub> | Mean | CI<sub>95%</sub> | Mean | CI<sub>95%</sub> |
| Pain Intensity | Delta | 4.8 | [-2.9, 12.4] | 14 | [6.9, 21.1] | 10.7 | [2.5, 19] | 9.4 | [-1.1, 19.9] |
| Pain Intensity | ANCOVA | 8.6 | [1.4, 15.7] | 11.8 | [5.2, 18.4] | 9.6 | [2.1, 17.1] | 8.9 | [-0.7, 18.5] |
| Pain Disability | Delta | 5.1 | [-5.2, 15.4] | 12.6 | [3, 22.3] | 10.2 | [-0.9, 21.3] | 7 | [-7.3, 21.2] |
| Pain Disability | ANCOVA | 9.9 | [0.1, 19.7] | 9.8 | [0.8, 18.8] | 8.7 | [-1.5, 19] | 6.4 | [-6.7, 19.4] |

Stratification by CPG grades shows, that the low pain group (grade 1, n = 74) did not improve on average in either pain intensity (2.0 CI_95_% [-1.7, 5.8]) or pain disability (−0.4 CI_95_% [-4.8, 3.9]). The longitudinal change was, thus, largely driven by the intermediate/high pain groups (grade 2, n = 39) with effect sizes of 18.2 CI_95_% [13.1, 23.3] for pain intensity and 13.4 CI_95_% [7.5, 19.4] for pain disability.

### Within-participant mediation – how does the treatment work?

Within the intervention group, changes in depression and hopelessness seem to be substantially associated with changes in pain intensity and pain disability (Figure 5). The indirect effect of depression on pain intensity was -2.2 HPDI_90_% [-4.4, -0.6] and on pain disability -1.9 HPDI_90_% [-4.3 -0.1]. The sum of indirect effects was -3.5 HPDI_90_% [-6.4, -0.8] for pain intensity and -4.7 HPDI_90_% [-8.6, -0.9] for pain disability, indicating potential treatment mechanisms by the supposed mediators The total proportion mediated was 34.1 % for pain and 49.3 % for disability.

**Figure 5.**
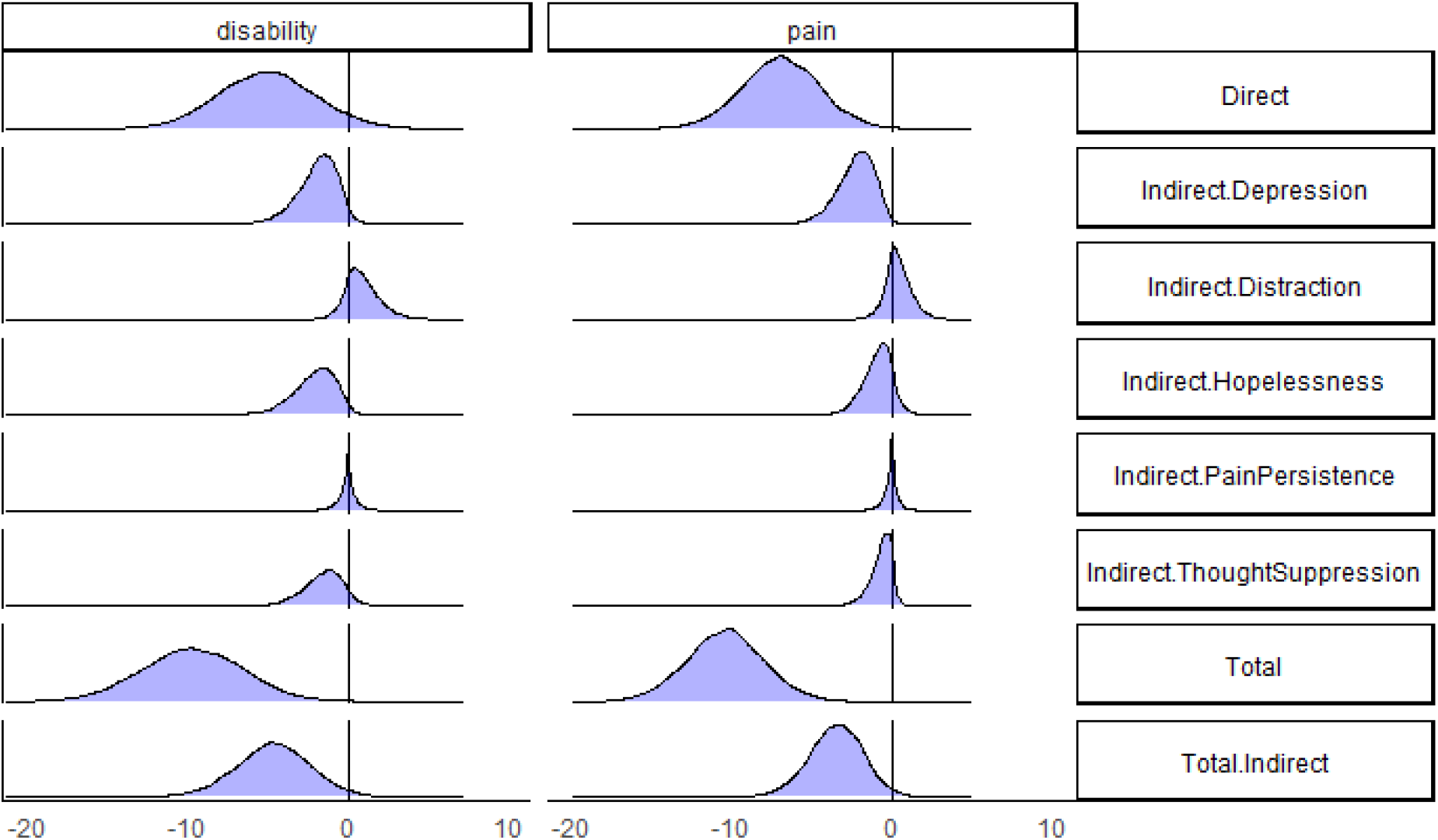
Posterior distributions for the direct, indirect and total (indirect) effects of the within-participant, (serial) multiple mediation model with two outcomes (pain intensity and pain disability).

## Discussion

We found similar decreases in the multimodal intervention group and the matched control groups for characteristic pain intensity and subjective pain disability. However, small treatment effects for strength and balance were in favor of the intervention. Distress-endurance subgroups showed the highest pain intensity and pain disability scores and subsequently, the highest reductions upon completion of the intervention.

### Population and sample representativeness

The intervention group consists of a heterogeneous sample. The classification approach via pain persistence and the BDI-PC yielded 32% distress-endurance responses, which is between study results from a subacute (19%) [16] and a chronic population (34 %) [12]. About one third of the intervention group showed high grades of chronification (Korff grades 2 and 3) with high pain and disability. Those people would rather seek care in the rehabilitative or tertiary care sector. Consequently, two thirds rather experience mild symptoms.

Nevertheless, the concept of the health care program we evaluated in this study aims at prevention, anchored in German social law (SGB V $20a). In our experience, this incongruency is common in practice, as minimal exclusion criteria exist. Consequently, practitioners are challenged by heterogenous groups, comprising the overall quality of such programs which is why the national guidelines suggest an early screening of affected people [23]. The avoidance-endurance model and other classification systems [21] as well as valid diagnostics such as the Keele StarT-Back [19] or RSI [47] might help to assign adequate treatments.

### Treatment effects

Interpreting the ATT for characteristic pain intensity and subjective pain disability, the finding of a “null” effect might surprise at first. In fact, both groups experienced reductions in pain intensity and disability. One may suspect that the effects of an intervention applied directly in health care might yield on average inferior results to a strictly controlled RCT [11]. Examples of such discrepancies between *efficacy* and *effectiveness in* the health care sector are already given by Cochrane (1972) [7] and Treweek & Zwarenstein [44]. The control group reduced disability by a fair amount (4.9 points) [31], which may be a result of selection bias (e.g. participants of back pain studies are likely to suffer recently from back pain and get better soon without treatment). Lastly, the average effect is likely to be comprised by the symptom heterogeneity in the intervention group, where most participants experienced rather mild symptoms.

The treatment effects for postural control were surprisingly high. Unpublished results from the multi-center RCT [32] show rather negligible, non-significant effects. In our study, the comparison with a matched control group from the same study yielded larger effect sizes for balance. The effects for flexion and extension were rather small but uncertain. We suppose, that this can be partly explained by stronger functional deficits in the intervention group indicated by higher age even after matching. For such people, the sensorimotor exercise may provide a much higher stimulus for adaptation, especially in the early phase of training. However, low baseline values also may yield larger effect sizes due to regression to the mean [41].

### Clinical relevance: For whom is the treatment effective?

The multimodal treatment might be most effective for distress-endurance response types, which are classified by high depression and high pain persistence scales. This result is in line with Cane et al. [6], who suggested, that changes toward adaptive responses (activity pacing) yield functional outcomes. A reduction in distress-endurance responses from pre to post was also observed in our study (22 to 12). In their Cochrane review, Kamper et al. [22] found medium effect sizes for multidisciplinary treatment in chronic low back pain populations for pain (d=0.55) and disability (d=0.41) reductions after 12 weeks of intervention. It seems reasonable to allocate multidisciplinary treatment to those with significant psychosocial impact [22].

Sensorimotor exercise might be particularly good for learning activity pacing to overcome the endurance pattern. This kind of exercise promotes physical activity by coordinative/balance training with trunk stability exercise with low demands on physical strength which was shown to be effective in reducing pain and disability [31]. This way, endurance response types learn effective regulation. Further, the educational component fosters self-reflection which should enhance self-efficacy via a biopsychosocial pathway. The average reductions in pain persistence and depression in almost all subgroups except for the adaptive response types at baseline might also indicate the effectiveness for the distress-endurance response type. The fear-avoidance response type might also benefit from this kind of intervention. The semi-standardized progression with a gradually increasing level structure can help to build trust in a movement to overcome kinesophobia.

### How does the treatment work?

We found an association between changes in mood scales and changes in the outcomes. Murillo et al. [30] found that depression is a commonly used mediator in mediation analysis. They also found that the mediated proportion for cognitive behavioral treatment did not exceed 20% for any mediator. The (total) mediated proportion in this study is about twice as large, which could be explained by the within-mediation model without a control group. Surprisingly, the smallest indirect effect was found for pain persistence. Pain persistence was a crucial scale for classifying AEM subgroups in several studies [16, 42, 48]. In conclusion, the average decrease in pain persistence seems to be a minor driver for changes in pain and disability than mood characteristics.

Contrary to this analysis, Liew et al. [27] suggested based on their data-driven analysis, that changes in depression might be affected by earlier changes in pain and disability. This switch between mediator and outcome yields the same model fit at least in a 3-parameter model (e.g., exposure, mediator, outcome). Therefore, based on only the data we cannot conclude whether pain and disability are reduced because of reduced depression/mood symptoms or the other way round. In another multicenter study from the MiSpEx-Network it was shown, that psychosocial factors are relevant moderators of the efficacy of sensorimotor exercise in people with unspecific low back pain. This was shown for the moderation of exercise effects on pain intensity due to depressive mood, vital exhaustion and perceived social support as well as for pain disability due to depressive mood, social satisfaction and anxiety. Contrary, there was no mediation of a psychosocial variable on the exercise effects on pain [46].

Further research is highly needed, as in general, treatment mechanisms for chronic pain are scarcely investigated and heterogeneous in most cases [30]. However, it becomes again more clear, that early identification of psychosocial risk factors by diagnostic tools, may be important for success in therapy.

### Assumptions for calculating the ATT

The ATT (Average Treatment Effect on Treated) can be calculated using matching based on certain assumptions. One is *common support* (also called positivity or overlap) of the matching covariates, which was reasonably indicated considering the balance after matching. More critique can be placed on the assumption of *conditional exchangibility* (also called conditional independence or strong ignorability), which is for the ATT, that the treatment assignment is independent of the potential (unseen) outcome for receiving no treatment when adjusting for a set of covariates [14, 35]. It can be questioned, whether the chosen covariates (baseline parameters, sex, age) are sufficient to satisfy this assumption. Further, the choice of the comparator influences the ATT estimate. The control group actively seeked treatment in form of study participation and received a more intense and frequent diagnostic including reminders to improve study adherence. Summarising these critiques, restrictions have to be placed on the interpretation of the ATT.

### Limitations

Due to the characteristics of this study (pragmatic trial), the drop-out was high and even higher among participants with high depression scores. One reason for the high drop-out rate was, missing direct contact with participants, i.e. the questionnaires were handed over by intermediaries like the health insurance and the coaches. In addition, the matched control group received a more extensive assessment and participated in a follow-up at 3 weeks but the intervention group did not. Furthermore, the intervention was not strictly controlled, leading to heterogeneity in delivery and, consequently, participant adherence. This heterogeneity was directly observed as feedback from the coaches who conducted the intervention. To overcome some of these issues, we carried out an extensive procedure (sequential matching analysis with sensitivity analysis), but the validity of the comparison might be questioned in light of the mentioned contextual factors. Further, the control group could not be used for moderation analysis, so we could not test the 3-fold interaction (time x intervention x AEM) but only a 2-fold interaction (time x AEM). Given that the derived AEM subgroup classifications are “natural” occurring groups rather than randomized, the coefficients from the baseline-adjusted moderation analyses are also biased estimators and do not yield causal estimates due to regression to the mean [41, 45]. Further, the sample size for the functional outcomes (n=18) is considerably low which might have led to inflated effect sizes.

## Conclusion

Perturbation exercise with education yielded only small treatment effects in a heterogenous population with rather mild symptoms. While restrictions in the interpretation have to be placed regarding the comparability, a pragmatic RCT should be conducted to evaluate the effectiveness. We found some evidence, that targeting depressive-endurance subgroups with a multimodal treatment approach can have an influence on treatment effects and should be considered in treatment planning and tailoring.

## Data Availability

Data of the treatment group, study documents and the analysis script are available in our online repository.

https://osf.io/mbrh

## Funding

This study was conducted within the MiSpEx research network and funded by the German Federal Institute for Sport Science (BISp) [ZMVI1-080102A/11-18].

## Declaration of interest

The authors declare no conflict of interest.

## Data availability

Data from the treatment group, study documents and the analysis script are available in our online repository: https://osf.io/mbrh9/

## Contributions

Conceptualization – RS, DF, TT, PP, Methodology – RS; DF, TT, MH, DN, Formal Analysis – RS, MH, Investigation – RS; TT, Resources – RS; PP, FM, CS, HB, DN MS, Data Curation – RS; TT, Writing: Original Draft – RS, DN, Writing: Review & Editing – All, Visualization – RS, Project administration & Funding – FM, PP

## Acknowledgments

We like to thank the participants. Also, we like to thank the health insurance AOK Nordwest for their help in the certification and implementation of the program, organizing the workshops for coaches, financial support for designing a course booklet and collecting paper-back questionnaires from the course participants. We also like to thank everyone who participated in data collection but did not meet the criteria for authorship.

## Notes

### Competing Interest Statement

The authors have declared no competing interest.

### Clinical Trial

Intervention: DRKS00030389, Control: DRKS00010129

### Author Declarations

The study was approved by the local ethics committee of the Faculty of Sport Science of Ruhr-University Bochum (EKS V 10/2017).

### Summary of Updates

Revisions based on reviewer and author feedback. - new matching and estimation of treatment effects - added results on matching success (balance) and discussion of matching assumptioms - clarification and structure

